# Metformin is Associated with Reduced COVID-19 Severity in Patients with Prediabetes

**DOI:** 10.1101/2022.08.29.22279355

**Authors:** Lauren E. Chan, Elena Casiraghi, Bryan Laraway, Ben Coleman, Hannah Blau, Adnin Zaman, Nomi Harris, Kenneth Wilkins, Michael Gargano, Giorgio Valentini, David Sahner, Melissa Haendel, Peter N. Robinson, Carolyn Bramante, Justin Reese, the N3C consortium

## Abstract

**Background:** With the continuing COVID-19 pandemic, identifying medications that improve COVID-19 outcomes is crucial. Studies suggest that use of metformin, an oral antihyperglycemic, is associated with reduced COVID-19 severity in individuals with diabetes compared to other antihyperglycemic medications. Some patients without diabetes, including those with polycystic ovary syndrome (PCOS) and prediabetes, are prescribed metformin for off-label use, which provides an opportunity to further investigate the effect of metformin on COVID-19.

**Participants:** In this observational, retrospective analysis, we leveraged the harmonized electronic health record data from 53 hospitals to construct cohorts of COVID-19 positive, metformin users without diabetes and propensity-weighted control users of levothyroxine (a medication for hypothyroidism that is not known to affect COVID-19 outcome) who had either PCOS (n = 282) or prediabetes (n = 3136). The primary outcome of interest was COVID-19 severity, which was classified as: mild, mild ED (emergency department), moderate, severe, or mortality/hospice.

**Results:** In the prediabetes cohort, metformin use was associated with a lower rate of COVID-19 with severity of mild ED or worse (OR: 0.630, 95% CI 0.450 - 0.882, p < 0.05) and a lower rate of COVID-19 with severity of moderate or worse (OR: 0.490, 95% CI 0.336 - 0.715, p < 0.001). In patients with PCOS, we found no significant association between metformin use and COVID-19 severity, although the number of patients was relatively small.

**Conclusions:** Metformin was associated with less severe COVID-19 in patients with prediabetes, as seen in previous studies of patients with diabetes. This is an important finding, since prediabetes affects between 19 and 38% of the US population, and COVID-19 is an ongoing public health emergency. Further observational and prospective studies will clarify the relationship between metformin and COVID-19 severity in patients with prediabetes, and whether metformin usage may reduce COVID-19 severity.

## Introduction

Since the onset of the coronavirus disease 2019 (COVID-19) pandemic caused by the severe acute respiratory syndrome (SARS)-associated coronavirus-2 (SARS-CoV-2), clinicians and researchers have sought new therapeutic options for COVID-19 patients. Current treatments, consisting of antiviral and immune-based interventions, have reduced but not eliminated COVID-19-related morbidity and mortality^1,2^. One strategy for identifying new agents is drug repurposing: identifying previously approved medications that may be effective for treatment of COVID-19. This process reduces the time and cost required to identify new therapeutic options compared to novel drug discovery.^3,4^

Several studies have proposed that metformin, a routinely prescribed antihyperglycemic agent, reduces COVID-19 severity in patients with diabetes taking the medication prior to onset of infection.^5–8^ Metformin decreases hepatic glucose production and improves insulin sensitivity by increasing peripheral glucose uptake and utilization.^9^ Metformin activity stimulates AMP-activated protein kinase (AMPK) in hepatocytes, which thereby reduces hepatic gluconeogenesis.^10^ Additional proposed mechanisms of metformin include inhibition of glucose-6-phosphate (G6P) leading to reduced hepatic gluconeogenesis, as well as decreased reactive oxygen species production at the respiratory-chain complex 1.^11,12^

Appropriate blood glucose management with metformin in COVID-19 patients prior to infection is associated with milder disease course, reduced incidence of acute respiratory distress syndrome (ARDS), and fewer cases of invasive care such as ventilator support.^13^ It has been suggested that metformin use may also reduce susceptibility to SARS-CoV-2 infection by increasing AMPK cell signaling, inhibiting binding of the viral spike protein, and preventing viral entry into the cell.^13,14^ Metformin may attenuate immune inflammatory responses and release of proinflammatory cytokines.^13^ As COVID-19 can present with cytokine storm, immune modulatory activity is desirable to avoid excessive inflammation and impaired organ function.^15^

Metformin is primarily prescribed for individuals with Type 2 Diabetes Mellitus (T2DM), but it is also recommended for off-label use in other groups. Two sizable populations who may benefit from off-label metformin usage are individuals who have prediabetes and those with polycystic ovary syndrome (PCOS).

Prediabetes is a state of intermediate hyperglycemia due to insulin resistance that poses a high risk for progression to diabetes. Individuals with prediabetes usually have a higher than normal variability of blood glucose concentration, but less than the threshold for diagnosing diabetes.^16^ The American Diabetes Association (ADA) 2021 guidelines define prediabetes as the presence of any the following: a fasting plasma glucose of 100 mg/dl to 125 mg/dl (5.6-6.9 mmol/L), a 2-hour plasma glucose during a 75-gram oral glucose tolerance test between 140 mg/dl to 199 mg/dl (7.8-11 mmol/L), or a hemoglobin A1c of 5.7% to 6.4% (39-47 mmol/mol).^17^ Without proper management between 25% to 50% of people with prediabetes progress to diabetes.^18^ Management for prediabetes involves intensive lifestyle changes, including weight loss, to reduce the risk of progression to diabetes. In addition, metformin can reduce or delay the incidence of diabetes in individuals with prediabetes.^19^

PCOS occurs in up to 12% of reproductive-aged women worldwide and is a diagnosis of exclusion made by the presence of 2 of the 3 Rotterdam Criteria (oligomenorrhea/amenorrhea, clinical or biochemical evidence of hyperandrogenism, and polycystic morphology of ovaries on ultrasound^20,21^) in absence of thyroid disorders, states of prolactin excess, and congenital adrenal hyperplasia^22^. Although insulin resistance is not a defining feature of PCOS, it is present in approximately75% of patients with PCOS independent of their BMI.^23,24^ However, the presence of obesity has been shown to reduce insulin sensitivity twofold, which in turn is thought to worsen the hyperandrogenism characteristic of PCOS.^24–26^ Despite extensive study, the mechanisms of impaired glucose utilization in PCOS remain unknown. There are no medications approved by the Food and Drug Administration (FDA) for PCOS and available therapies are largely meant to manage symptoms.^27^ One of these therapies is metformin, which is a commonly used off-label in patients with PCOS to increase insulin sensitization, induce ovulation, regulate menstrual cycles, and aid weight loss.^23^

For individuals at high risk for severe COVID-19, therapeutic approaches to prophylactically prevent poor COVID-19 outcomes are highly desirable. Given that many patients with prediabetes and PCOS are both high risk and already take metformin, they are an ideal population for evaluating the impacts of metformin usage prior to COVID-19 infection onset. We hypothesize that documented usage of metformin prior to COVID-19 infection will be associated with decreased severity of COVID-19 infection outcomes. In turn, metformin use prior to infection may be an affordable way to improve COVID-19 outcomes for patients with prediabetes via glycemic control and other mechanisms.

## Methods

This retrospective observational study utilized clinical patient data aggregated in the National COVID Cohort Collaborative (N3C) (covid.cd2h.org). The N3C Data Enclave harmonizes electronic health record (EHR) data from over 13 million patients in the United States from a total of 74 institutions. This investigation leveraged data from 53 institutions that provided cases who met the inclusion criteria. All data and software used in the present study are available within the N3C Data Enclave (covid.cd2h.org).

Data within the N3C enclave is harmonized from source clinical data models into Observational Medical Outcomes Partnership (OMOP)^28^ version 5.3.1 format. The OMOP model includes standardized definitions of conditions, lab tests, procedures, and other relevant clinical data including positive COVID-19 laboratory tests.^29,30^

### Definition of metformin and levothyroxine use

We defined metformin users as those patients who had recorded use^31^ of metformin beginning on or before the start date of the visit during which the patient was diagnosed with COVID-19 and overlapping for at least one day with the COVID-19 visit. We also identified use of levothyroxine within our study population as a comparator drug. As with metformin, individuals were defined as using levothyroxine if they had a drug era for levothyroxine beginning on or before the date of COVID-19 diagnosis and continuing for at least one day. Any patients using both levothyroxine and metformin were excluded from the study population (n < 20 for PCOS, n =38 for prediabetes).

### PCOS and Prediabetes Cohorts

Our study included patients who were identified as being COVID-19 positive by positive SARS-CoV-2 laboratory test (polymerase chain reaction or antigen) after January 1, 2020. Positive patients were assigned to our two cohorts of prediabetes and patients with PCOS as follows. Patients were identified as having prediabetes if they had either a documented history of the condition of prediabetes (Supplement 1) or hemoglobin A1C (HbA1C) between 5.7% and 6.4% (American Diabetes Association-recognized range for prediabetes)^32^. Patients were identified as having PCOS if they had a documented history of the condition (Supplement 1).

We removed patients from both cohorts who 1) had a documented history of the condition of diabetes mellitus type 1, diabetes mellitus type 2, gestational diabetes, or another diagnosed condition that is characterized by abnormal blood glucose (Supplement 1); or 2) a documented HbA1C) ≥ 6.5% (American Diabetes Association-recognized range for diabetes).^32^

OMOP concept ID codes for all conditions and medications used in this analysis are listed in Supplement 1. Codesets containing the relevant concept IDs for each construct were formulated using ATLAS (http://atlas-covid19.ohdsi.org/), the graphical user interface for the OMOP common data model.^33^

### Study Design

We explored the potential association between treatment with metformin and severity of COVID-19 within each of the cohorts. To further correct residual covariate imbalance within the cohorts, we performed inverse probability weighting.

### Collinearity, missingness imputation, and statistical analysis

Covariates were combined or removed when significant collinearity was detected. Collinearity between covariates was assessed by calculating the generalized variance inflation factor (GVIF).^34,35^ We observed high GVIF for race and Charlson Comorbidity Score (CCI)^36^ (data not shown), likely due to low representation in a variety of race categories and CCI values. To alleviate that collinearity, while still including race and CCI within the regression, the race category was modified into a binary “white”/“non-white” variable, and the CCI variable was converted to a binary variable “CCI ≤1”/“CCI >1”. No other included variables showed a GVIF greater than 5.

Missing values were imputed for BMI, race (white versus non-white), and ethnicity. We compared various imputation models including missRanger^37,38^ and different MICE imputers^39^ on a (limited) complete dataset. All the models resulted in comparable estimates and we therefore chose the missForest implementation provided by missRanger (with a number of trees in the random forest equal to 101) because our experiments on a larger patient cohort showed its higher validity when compared to the MICE models.^40^

Inverse probability weighting was performed using the “`Weightthem`” method implemented in the R MatchThem package^41^ using “`propensity score`” weights, the Average Treatment Effect on the Treated (`ATT`) estimand, and the “`within`” approach. Propensity score weighting was performed within each imputed dataset, all the weighted datasets are input to individual logistic regression models and the obtained estimates were then pooled via Rubin’s rule.^42^ As such, for each imputed dataset, patients within a cohort (e.g. prediabetes, PCOS) with recorded metformin use were weighted to improve the overall matching with respect to another patient cohort with recorded levothyroxine but not metformin use. The propensity score formula included age, race, ethnicity, sex (only included for prediabetes), smoking status or nicotine dependence, Charlson Comorbidity Index, BMI, and prediabetes status (only included for PCOS), PCOS status (only included for prediabetes) and a set of comorbidities with prevalence higher than 2% (see Table 1).

**Table 1A.**
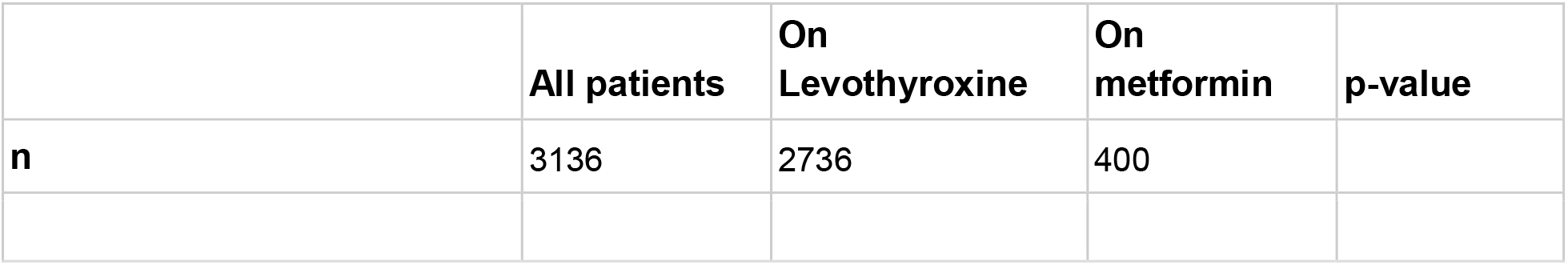
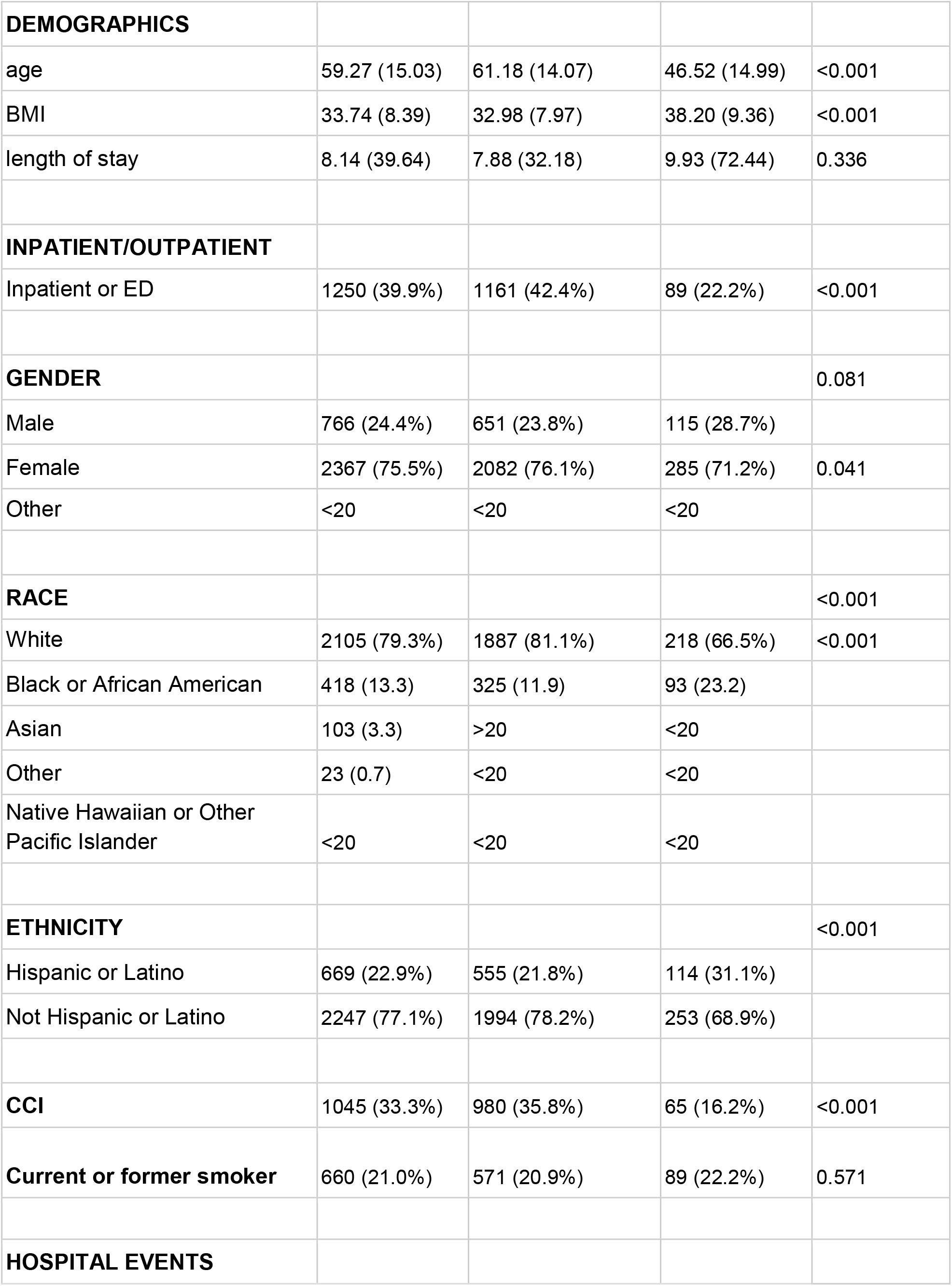
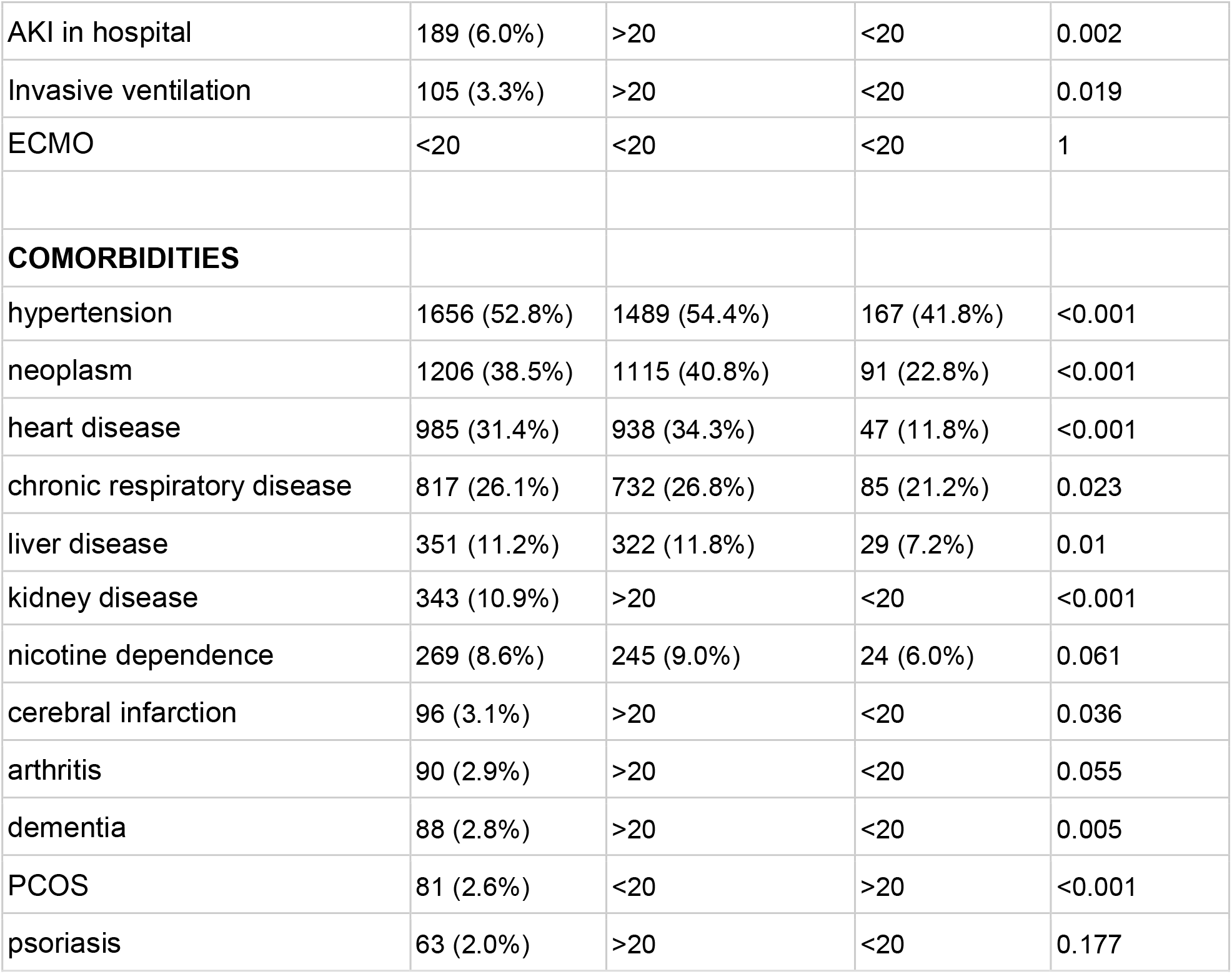
Demographics of the prediabetes cohort

|  | All patients | On<br>Levothyroxine | On<br>metformin | p-value |
| --- | --- | --- | --- | --- |
| **n** | 3136 | 2736 | 400 |  |
| **DEMOGRAPHICS** |  |  |  |  |
| age | 59.27 (15.03) | 61.18 (14.07) | 46.52 (14.99) | <0.001 |
| BMI | 33.74 (8.39) | 32.98 (7.97) | 38.20 (9.36) | <0.001 |
| length of stay | 8.14 (39.64) | 7.88 (32.18) | 9.93 (72.44) | 0.336 |
| **INPATIENT/OUTPATIENT** |  |  |  |  |
| Inpatient or ED | 1250 (39.9%) | 1161 (42.4%) | 89 (22.2%) | <0.001 |
| **GENDER** |  |  |  | 0.081 |
| Male | 766 (24.4%) | 651 (23.8%) | 115 (28.7%) |  |
| Female | 2367 (75.5%) | 2082 (76.1%) | 285 (71.2%) | 0.041 |
| Other | <20 | <20 | <20 |  |
| **RACE** |  |  |  | <0.001 |
| White | 2105 (79.3%) | 1887 (81.1%) | 218 (66.5%) | <0.001 |
| Black or African American | 418 (13.3) | 325 (11.9) | 93 (23.2) |  |
| Asian | 103 (3.3) | >20 | <20 |  |
| Other | 23 (0.7) | <20 | <20 |  |
| Native Hawaiian or Other Pacific Islander | <20 | <20 | <20 |  |
| **ETHNICITY** |  |  |  | <0.001 |
| Hispanic or Latino | 669 (22.9%) | 555 (21.8%) | 114 (31.1%) |  |
| Not Hispanic or Latino | 2247 (77.1%) | 1994 (78.2%) | 253 (68.9%) |  |
| **CCI** | 1045 (33.3%) | 980 (35.8%) | 65 (16.2%) | <0.001 |
| **Current or former smoker** | 660 (21.0%) | 571 (20.9%) | 89 (22.2%) | 0.571 |
| **HOSPITAL EVENTS** |  |  |  |  |
| AKI in hospital | 189 (6.0%) | >20 | <20 | 0.002 |
| Invasive ventilation | 105 (3.3%) | >20 | <20 | 0.019 |
| ECMO | <20 | <20 | <20 | 1 |
| **COMORBIDITIES** |  |  |  |  |
| hypertension | 1656 (52.8%) | 1489 (54.4%) | 167 (41.8%) | <0.001 |
| neoplasm | 1206 (38.5%) | 1115 (40.8%) | 91 (22.8%) | <0.001 |
| heart disease | 985 (31.4%) | 938 (34.3%) | 47 (11.8%) | <0.001 |
| chronic respiratory disease | 817 (26.1%) | 732 (26.8%) | 85 (21.2%) | 0.023 |
| liver disease | 351 (11.2%) | 322 (11.8%) | 29 (7.2%) | 0.01 |
| kidney disease | 343 (10.9%) | >20 | <20 | <0.001 |
| nicotine dependence | 269 (8.6%) | 245 (9.0%) | 24 (6.0%) | 0.061 |
| cerebral infarction | 96 (3.1%) | >20 | <20 | 0.036 |
| arthritis | 90 (2.9%) | >20 | <20 | 0.055 |
| dementia | 88 (2.8%) | >20 | <20 | 0.005 |
| PCOS | 81 (2.6%) | <20 | >20 | <0.001 |
| psoriasis | 63 (2.0%) | >20 | <20 | 0.177 |

To investigate the association of metformin and other covariates with COVID-19 severity, we applied the `glm` function of the stats R package^43^ to perform logistic regression on each imputed and matched dataset.

COVID-19 severity was defined using the ordinal levels of “mild”, “mild ED”, “moderate”, “severe”, and “mortality/hospice”^30^, and it was used to create four derived outcome variables based on COVID-19 severity: mild ED or worse, moderate or worse, severe or worse, and mortality/hospice. Each of outcome, we independently performed a logistic regression (LR) to evaluate COVID-19 severity and its association with metformin, including also the predictors of age, race, ethnicity, sex (only included for prediabetes), smoking status, Charlson Comorbidity Index, BMI, prediabetes (only included for PCOS), PCOS (only included for prediabetes) and comorbidities. Since in the PCOS cohort, the number of patients with COVID-19 severity of severe and mortality/Hospice patients in the PCOS cohort was very small, we omitted LR for these two outcome variables. For “on metformin” status, we pooled the LR estimates across all the imputed datasets by Rubin’s rule and we recorded the corresponding p-values, the pooled odds ratios and their 95% confidence intervals. The p-values obtained for the “on metformin” status on the prediabetes and PCOS cohorts were adjusted by Bonferroni correction to account for family-wise FDR.

We used the *EValue* R package (version 4.1.2) to determine the minimum strength of an unmeasured confounder in the logistic regression that would be required to change the conclusion that metformin was associated with reduced severity of COVID-19.^44^ We treated the outcome of decreased COVID-19 severity as a non-rare outcome (as it occurred more frequently than 15%). For a more conservative estimate, we report the E-value estimate for the confidence interval of the odds ratio that is closest to the null, as recommended by VanderWeele et al.

## Results

Within the current study, two cohorts of COVID-19 positive individuals were identified for assessing the association between metformin usage and COVID-19 severity outcomes. The two cohorts developed were prediabetes and PCOS. Data for this project was derived from 53 total sites, among which only 26 provided PCOS case data. For this study, data was included up to May 12, 2022. The predictors with missing data were race (17% in PCOS, 15% in prediabetes), ethnicity (10% in PCOS and 7% in prediabetes), and BMI (14% in PCOS, 24% in prediabetes). Overall, the percentage of cases with any missing values was 30% in PCOS and 38% in prediabetes patients (Little’s test supported the assumption of Missing At Random over Missing Completely at Random data - p-value < 0.05^45^).

We evaluated a total of 3337 patients (of which 81 were in common between the two cohorts) with COVID-19 in this retrospective study, identified by the filtering steps described in Figure 1 (3136 in the prediabetes cohort, 282 in the PCOS cohort). Table 1 shows the demographics of both cohorts, including the Charlson Comorbidity Index score.

**Figure 1.**
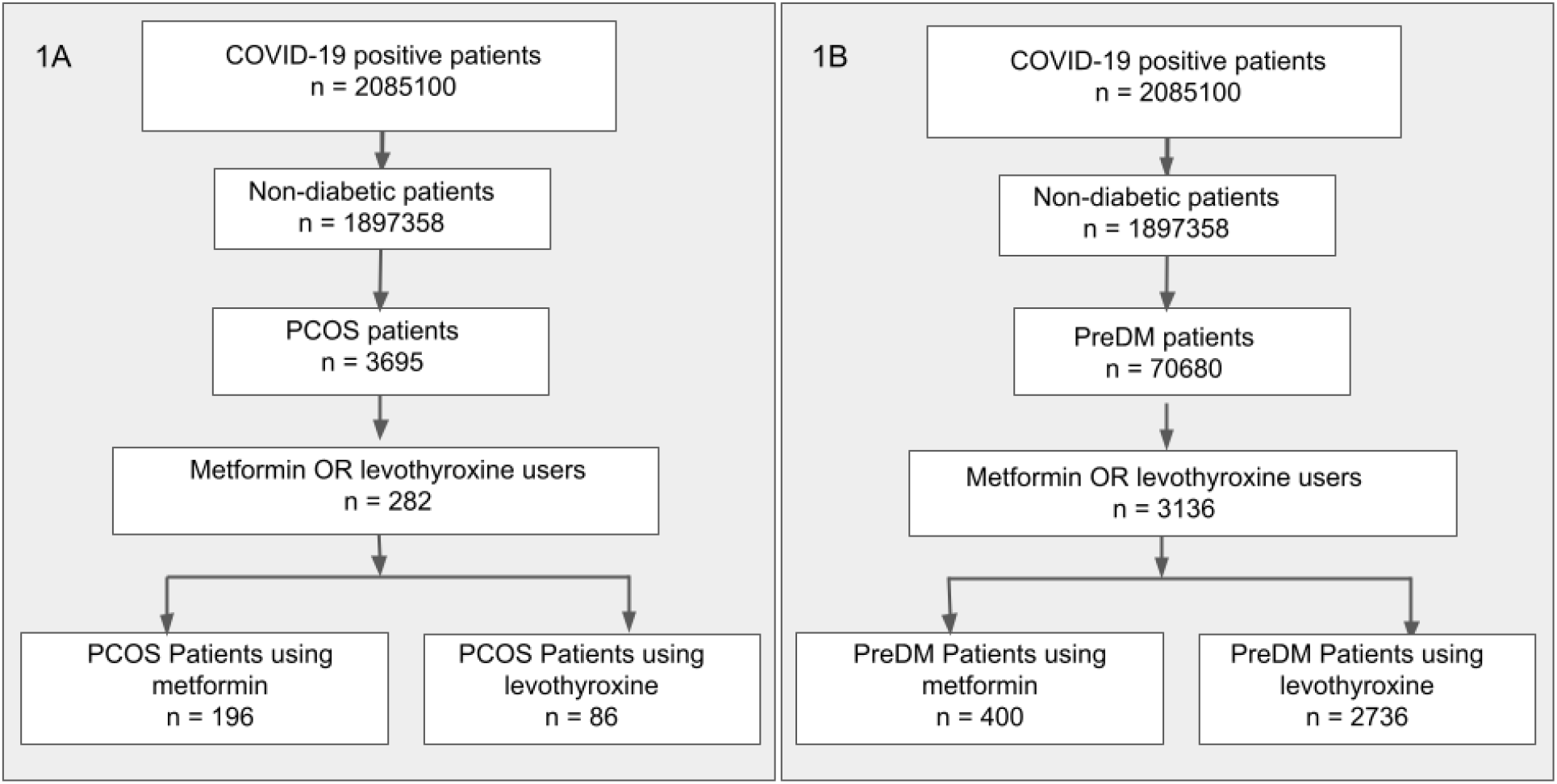
Definition of prediabetes and PCOS cohorts. COVID-19 positive patients were filtered to remove any individuals with diabetes and then separated into PCOS (1A) and prediabetes (1B) cohorts. For both cohorts, we selected patients that had a recorded usage of either metformin or levothyroxine.

**Table 1B.** Demographics of the PCOS cohort.

|  | **All patients** | **On**<br>**Levothyroxine** | **On metformin** | **p-value** |
| --- | --- | --- | --- | --- |
| **n** | 282 | 86 | 196 |  |
| **DEMOGRAPHICS** |  |  |  |  |
| age | 33.18 (8.42%) | 35.81 (8.06%) | 32.03 (8.33%) | <0.001 |
| BMI | 37.90 (8.94) | 36.73 (8.38) | 38.43 (9.15) | 0.173 |
| length of stay | 7.04 (39.95) | 12.51 (64.96) | 4.63 (21.00%) | 0.128 |
| **INPATIENT/OUTPATIENT** |  |  |  |  |
| Inpatient or ED | 72 (25.5%) | 28 (32.6%) | 44 (22.4%) | 0.1 |
| **RACE** |  |  |  | 0.379 |
| White | 190 (67.4) | 65 (75.6) | 125 (63.8) | 0.059 |
| Black or African American | 33 (11.7) | < 20 | > 20 |  |
| Asian | < 20 | < 20 | < 20 |  |
| Other | < 20 | < 20 | < 20 |  |
| Native Hawaiian or Other Pacific Islander | < 20 | < 20 | < 20 |  |
| **ETHNICITY** |  |  |  | 0.85 |
| Hispanic or Latino | 63 (24.8) | < 20 | >20 |  |
| Not Hispanic or Latino | 191 (75.2) | 59 (76.6) | 132 (74.6) |  |
| **CCI** | 30 (10.6) | < 20 | < 20 | 0.008 |
| **Current or former smoker** | 59 (20.9) | < 20 | >20 | 0.267 |
| **HOSPITAL EVENTS** |  |  |  |  |
| AKI in hospital | <20 | <20 | <20 | 0.671 |
| Invasive ventilation | <20 | <20 | <20 | 0.671 |
| ECMO | 0 | 0 | 0 | - |
| **COMORBIDITIES** |  |  |  |  |
| prediabetes | 84 (29.8%) | 18 (20.9%) | 66 (33.7%) | 0.044 |
| neoplasm | 57 (20.2) | 22 (25.6) | 35 (17.9) | 0.185 |
| hypertension | 54 (19.1) | < 20 | >20 | 0.992 |
| chronic respiratory disease | 49 (17.4) | < 20 | >20 | 1 |
| heart disease | 21 (7.4) | < 20 | < 20 | 0.302 |
| liver disease | < 20 | < 20 | < 20 | 0.005 |
| nicotine dependence | < 20 | < 20 | < 20 | 0.966 |
| kidney disease | < 20 | < 20 | < 20 | 0.759 |

### Outcomes

The outcome of interest for this investigation was COVID-19 clinical severity. Clinical severity was classified using the Clinical Progression Scale (CPS) established by the World Health Organization (WHO) for COVID-19 clinical research^46^ modified into the five following categories: “mild” (outpatient, WHO severity 1-3); “mild ED” (outpatient with ED visit, WHO severity 3); “moderate” (hospitalized without invasive ventilation, WHO severity 4-6); “severe” (hospitalized with invasive ventilation or ECMO, WHO severity 7-9); and “mortality/hospice” (hospital mortality or discharge to hospice, WHO Severity 10).^30^ To develop ordinal classifications for patient severity, patients were assigned to severity groups according to the maximum clinical severity during their index encounter^30^, which was defined as the medical encounter during which a positive COVID-19 test was documented for the first time. In Table 2 we report the COVID-19 severity distribution per cohort. The PCOS cohort did not include patients stratified across all five categories, but the prediabetes cohort spanned across all five categories of the severity scale.

**Table 2.** COVID-19 severity outcome of patients in prediabetes and PCOS cohorts. Values less than 20 patients are obscured to reduce the risk of patient reidentification. From the entire cohort of 3695 patients with PCOS, a total of 282 patients had recorded use of metformin or levothyroxine during COVID-19 presentation. Out of the entire cohort of 70680 patients with prediabetes, a total of 3136 had a recorded use of metformin or levothyroxine during COVID-19 presentation.

|  | PCOS |  |  | Prediabetes |  |  |
| --- | --- | --- | --- | --- | --- | --- |
|  | all | metformin | levothyroxine | all | metformin | levothyroxine |
| n | 282 | 196 | 86 | 3136 | 400 | 2736 |
| Mild | 210 (74.5%) | 152 (77.6%) | 58 (67.4%) | 1875 (59.8%) | 310 (77.5%) | 1565 (57.2%) |
| Mild ED | 34 (12.1%) | >20 | <20 | 191 (6.1%) | 31 (7.8%) | 160 (5.8%) |
| Moderate | 36 (12.8%) | >20 | <20 | 849 (27.1%) | 51 (12.8%) | 798 (29.2%) |
| Severe | < 20 | <20 | <20 | 57 (1.8%) | < 20 | > 20 |
| Mortality / hospice | < 20 | < 20 | <20 | 164 (5.2%) | < 20 | > 20 |

Within each cohort, the relationship between metformin use and other covariates (age, race, ethnicity, sex, smoking status, Charlson comorbidity, BMI, and comorbidities) and COVID-19 severity (“mild”, “mild ED”, “moderate”, “severe”, “mortality/hospice”) was evaluated. The control matched participants within the comparison are individuals with prediabetes who used levothyroxine. Levothyroxine is a synthetic thyroid hormone medication that can be used to treat hypothyroidism as well as other thyroid related conditions^47^. Similar to metformin, levothyroxine is a prescription only medication in the United States, is an oral medication, and requires daily usage and adherence for maximal therapeutic benefit. Levothyroxine is not known to be a successful therapeutic intervention for COVID-19, and has substantial usage in our study population. These considerations led to our choice of levothyroxine as an inactive comparator drug^48^ in this investigation.

Within the PCOS cohort, no significant association between metformin usage and decreased COVID-19 severity was observed (Figure 3). In the PCOS cohort, White race was significantly associated with COVID-19 severity in both of the logistic regressions, but no other significant associations were observed (Supplementary Figures S1a-b).

**Figure 2.**
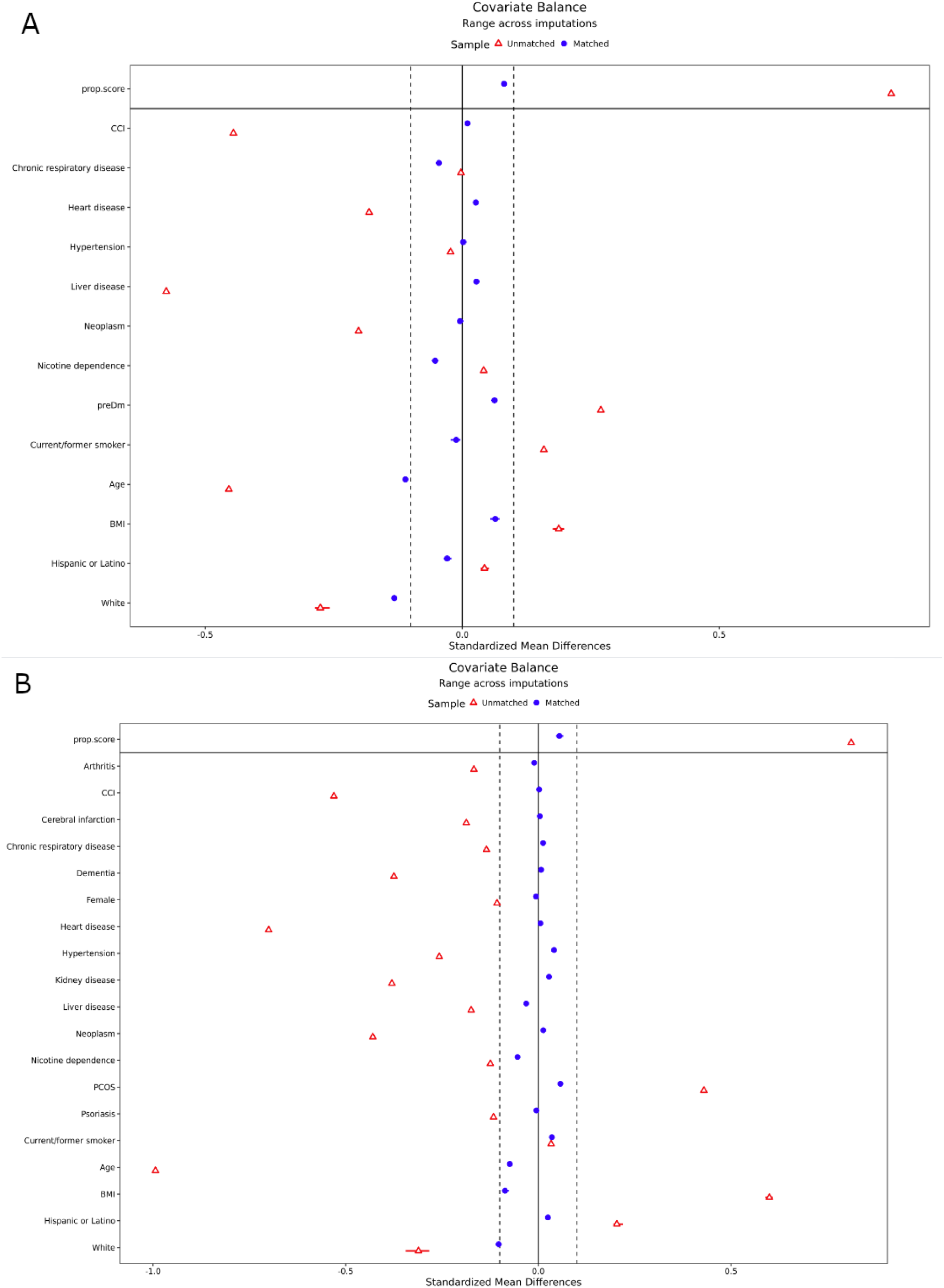
Inverse probability weighting in PCOS and prediabetes cohorts. Both PCOS (A) and prediabetes (B) cohorts were balanced using an inverse probability weighting approach. The standardized mean difference (x axis) is shown for each of 19 covariates (y axis) and the overall propensity score. Red triangles indicate the original covariate balance and blue circles indicate the covariate balance after weighting.

**Figure 3.**
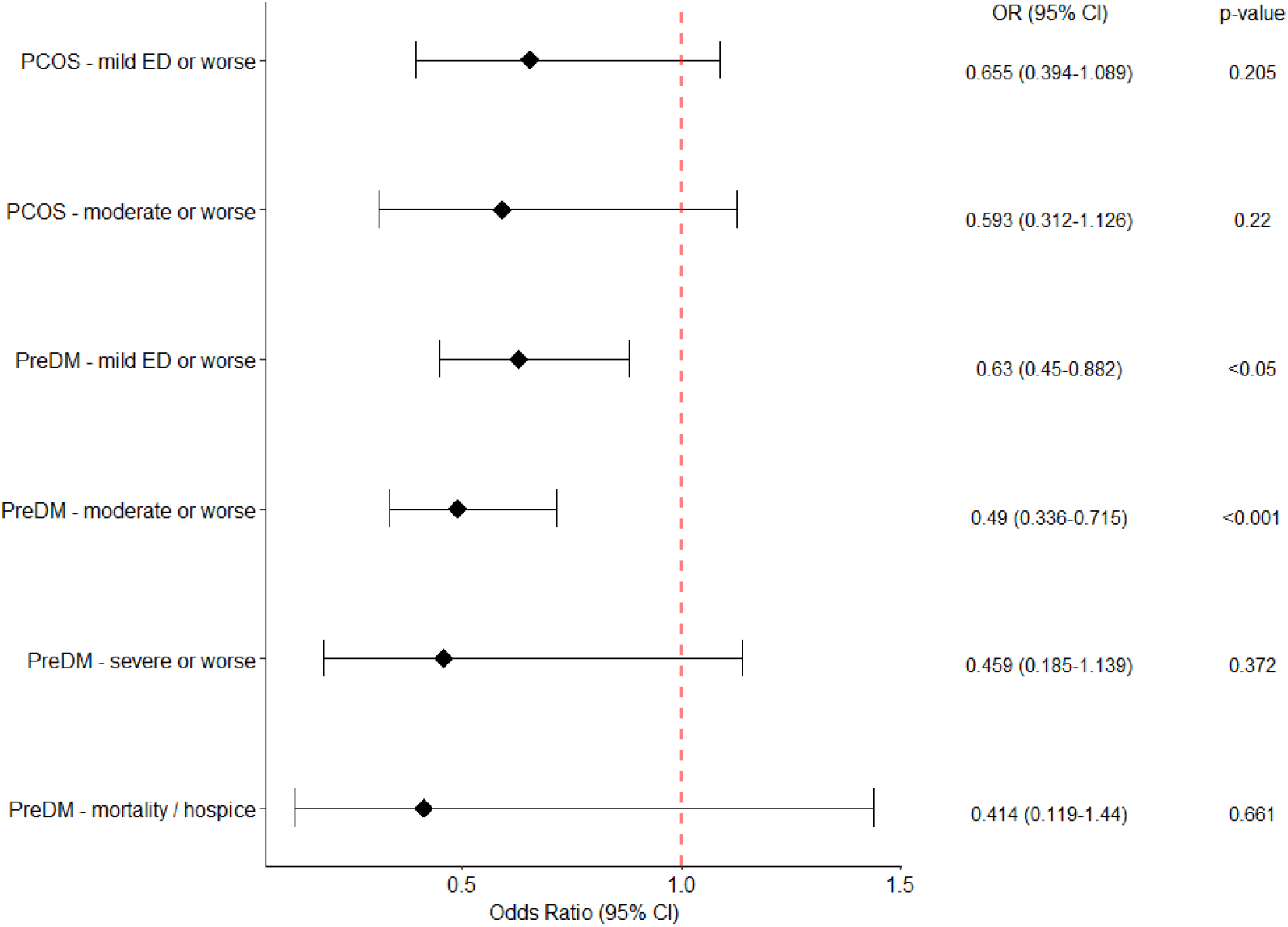
Association between metformin usage and COVID-19 severity in PCOS and prediabetes cohorts. †For the PCOS cohort, insufficient patients with severe or worse disease and mortality/hospice were available for a logistic regression to be performed. All p-values are Bonferonni corrected.

Assessments of the prediabetes cohort indicated a significant association of metformin usage with decreased incidence of COVID-19 severity of moderate or worse (Figure 3). In agreement with other prior investigations, we observed a significant association of COVID-19 severity moderate or worse with age for patients with prediabetes (Figure 4b). Unexpectedly, for patients with prediabetes we saw significantly decreased incidence of more severe COVID-19 in patients with chronic respiratory disease (Figures 4a and b), or hypertension (Figures 4a and b and supplementary Figures S2a) or for those with Hispanic or Latino ethnicity (Figures 4a and b), possibly due to the non-collapsibility of odds ratios^49,50^ and/or confounding factors such as behavior. A secondary analysis using logistic regression confirmed that metformin as well as Hispanic or Latino ethnicity, are associated with decreased incidence of more severe COVID-19; on the other hand, chronic respiratory disease and hypertension do not show any significant association with COVID-19 severity (Supplementary Figure S3). Heart disease (Figures S4a and 4b), and history of neoplasm (Supplementary Figure S1a) were variably associated with a higher incidence of severe disease outcomes.

**Figure 4.**
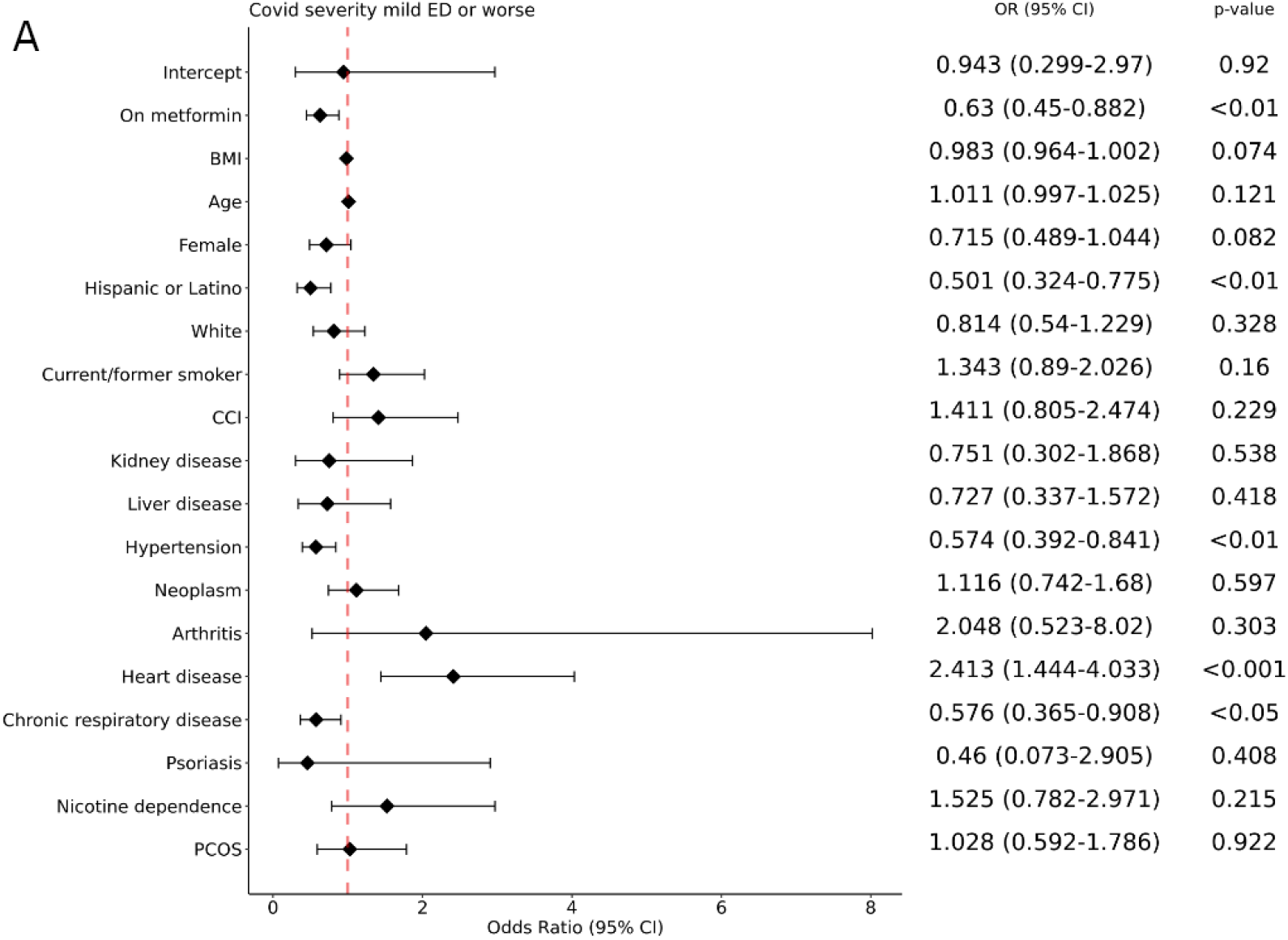
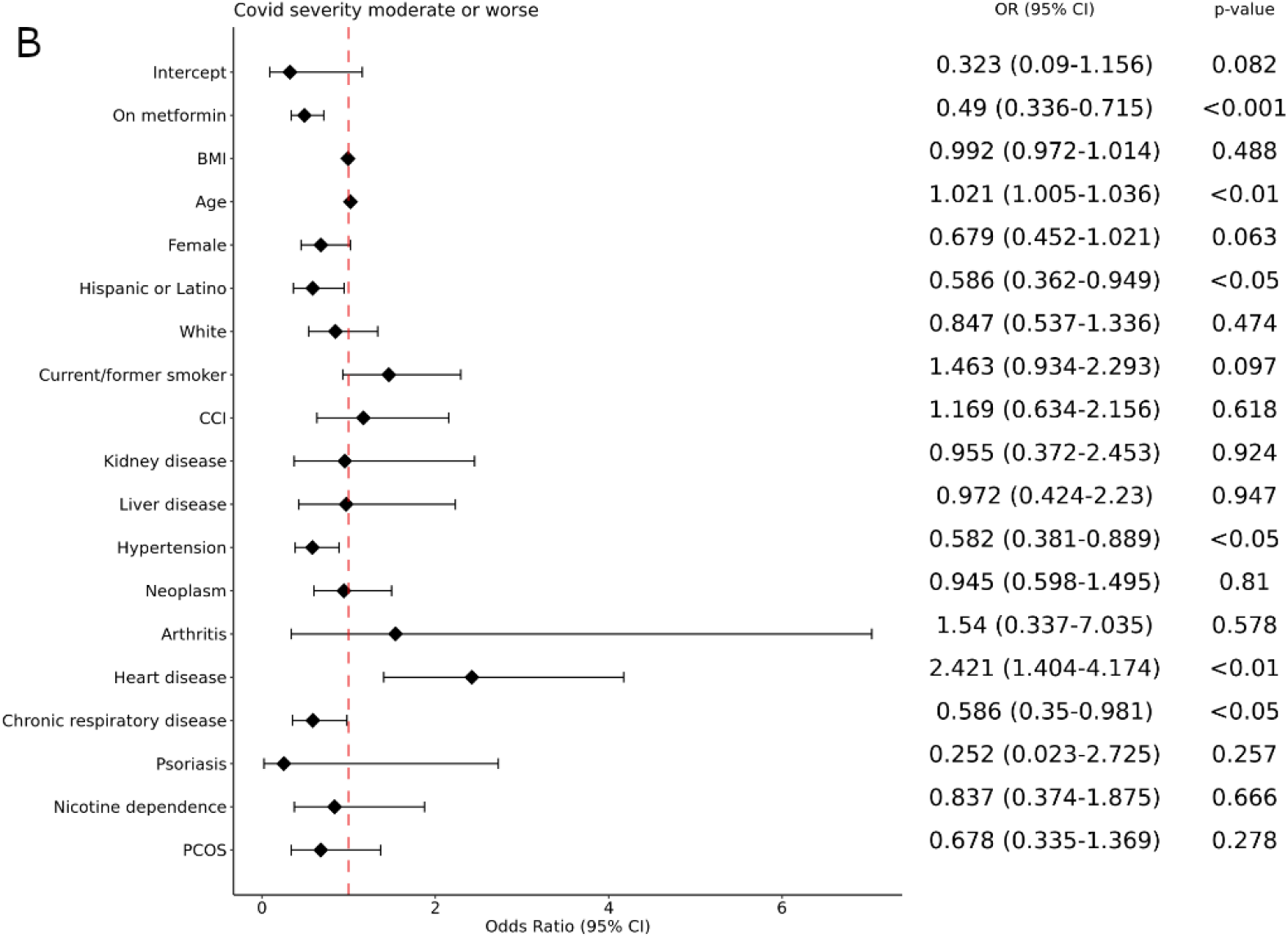
Forest Plots for Prediabetes. A: Mild vs Mild ED and worse, B: Mild and mild ED vs moderate and worse (Forest plots for comparisons of Mild, mild ED, and moderate vs severe and worse, and for Mild, mild ED, moderate, and severe vs death with Covid-19 are reported in Supplementary Figure S2).

We calculated the E-value for the observed values of the odds ratio to assess the sensitivity of our findings to uncorrected confounders.^44^ To explain the association of metformin use with decreased COVID-19 severity in the prediabetes cohort, an unmeasured confounder would need to be associated with both the treatment and the outcome with an odds ratio of 0.79 (upper bound 0.93) (mild ED or worse) and 0.70 (upper bound 0.84) (moderate or worse) above and beyond the confounders included in the regression.

## Discussion

We observed a significant association between metformin use and less severe COVID-19 in the prediabetes cohort compared with levothyroxine use. While multiple studies have been conducted to assess metformin impact on COVID-19 in the type 2 diabetes population, this is the largest study investigating off-label usage including individuals with prediabetes and PCOS. These findings agree with prior investigations in type 2 diabetes patients, in which metformin users were reported to have lower COVID-19 severity.^6,13,51^

Considering the impact of COVID-19 on populations investigated in this study, additional potential interventions to prevent or mitigate COVID-19 severity are greatly desired. While PCOS is a common endocrine disorder, the etiology and pathophysiology are not fully elucidated and clinical care for patients with PCOS can be inconsistent even during non-pandemic times.^52^ Additionally, a wide swath of risk factors of severe COVID-19 are also commonly observed in patients with PCOS, including obesity, hypertension, and metabolic syndrome.^52^ As these cardio-metabolic risk factors are more frequently seen in the patients with PCOS compared to the general population, patient risk for acquiring COVID-19 with severe presentation is increased.^53^ One report indicated that individuals with PCOS have a 28% increased risk of COVID-19 infection.^54^

The current study’s lack of association of metformin with COVID-19 outcome in the PCOS cohort may indicate that either metformin’s mechanism for reducing COVID-19 severity is not operative in patients with PCOS, or that our study was not sufficiently powered to detect the improvement mediated by metformin. Of some note, although not statistically significant, point estimates of the odds ratio (metformin versus levothyroxine) were directionally consistent in both populations. Given the potential impact an alternative COVID-19 therapy may pose for patients with PCOS, further investigation into this population with metformin and other drug repurposing candidates seems warranted.

Individuals with prediabetes also face increased COVID-19 risk compared to the general population. Although not meeting the threshold for type 2 diabetes diagnosis, individuals with prediabetes experience reduced insulin sensitivity and glycemic dysregulation.^55^ The consistent moderate hyperglycemia experienced by individuals with prediabetes can be accompanied by chronic vascular complications including blood pressure changes and cognitive dysfunction.^56^ Additionally, prediabetes may warrant consideration as a comorbidity independent of type 2 diabetes.^55^ From our investigation, metformin usage in individuals with prediabetes was associated with a significant decrease in COVID severity when compared to levothyroxine users using a binary categorization (mild + mild ED vs moderate and worse category). These findings support the association of metformin use with reduced COVID severity. Limited numbers of subjects with the most severe forms of disease and/or mortality did not enable us to assess the extent to which metformin lowers the risk of mechanical ventilation or death in patients with prediabetes. Future investigations using prospective trials and potentially randomized controlled trials are needed to elucidate whether metformin is associated with better COVID-19 outcomes.

### Limitations

This analysis is retrospective and is subject to confounding. Our study utilizes electronic health record data, which may contain some inaccuracies such as incomplete recording of patient data, and incomplete/incorrect translation of source EHR data to the OMOP format used by N3C, despite efforts to harmonize and quality-check these data centrally. Further, while metformin and levothyroxine prescriptions were assumed to be consistent with usage, medication compliance levels are not reported in our data set.

We chose levothyroxine as an inactive comparator drug because of its similarity to metformin with respect to typical daily usage requirement, prescription requirement, and primarily oral administration modality,^57^ and because levothyroxine is not known to affect COVID-19 severity. We were unable to select a similar oral antihyperglycemic medication (e.g. sulfonylureas, thiazolidinediones) for comparison with metformin due to limited usage of those medications within our study cohorts in the populations of interest.

We observed a limited number of patients (<20) in the severe and mortality/hospice category in both the PCOS and prediabetes cohorts, which may have limited the statistical power to detect an association between metformin use and COVID-19 severity. Future studies with greater cohort sizes may address this limitation.

It is possible that excessive or deficient levels of thyroid hormone in patients with thyroid disease may be associated with increased risk for poor COVID severity outcomes^58,59^, which may introduce a bias toward more severe COVID-19 outcome in patients using levothyroxine. This is contradicted in other investigations, particularly with thyroid diseases that are being actively managed.^60,61^

## Conclusion

In summary, this retrospective analysis of electronic health record data supports the previously described association between lower COVID-19 disease severity and metformin usage. However, the inherent limitations of observational analyses leave opportunities for future work in this area. The generalizability of our findings should be evaluated and future research utilizing a larger sample and/or a prospective design, including relevant immunologic biomarkers, may be beneficial to understanding the effect of metformin on COVID severity and any underlying mechanism.

## Supporting information

Supplement Tables - OMOP concept IDs

## Data Availability

Data and software used in the present study are available within the N3C Data Enclave (covid.cd2h.org).

http://covid.cd2h.org

## Funding

This work was supported by NCATS U24 TR002306. Additionally, Justin T. Reese was supported by the Director, Office of Science, Office of Basic Energy Sciences of the U.S. Department of Energy Contract No. DE-AC02-05CH11231; Adnin Zaman was supported by NIH NIDDK grant F32 DK123878. Melissa Haendel was supported by the Marsico Family at the University of Colorado Anschutz; Elena Casiraghi and Giorgio Valentini were supported by Università degli Studi di Milano, Piano di sviluppo di ricerca, grant 2015-17 PSR2015-17.

## Acknowledgements

The analyses described in this publication were conducted with data or tools accessed through the NCATS N3C Data Enclave covid.cd2h.org/enclave and supported by CD2H - The National COVID Cohort Collaborative (N3C) IDeA CTR Collaboration 3U24TR002306-04S2 NCATS U24 TR002306, DUR: RP-7BE1AC. This research was possible because of the patients whose information is included within the data from participating organizations (covid.cd2h.org/dtas) and the organizations and scientists (covid.cd2h.org/duas) who have contributed to the on-going development of this community resource ^29^. The content is solely the responsibility of the authors and does not necessarily represent the official views of the National Institutes of Health or the N3C program.

The N3C data transfer to NCATS is performed under a Johns Hopkins University Reliance Protocol #IRB00249128 or individual site agreements with NIH. The N3C Data Enclave is managed under the authority of the NIH; information can be found at https://ncats.nih.gov/n3c/resources.

We gratefully acknowledge the following core contributors to N3C: Adam B. Wilcox, Adam M. Lee, Alexis Graves, Alfred (Jerrod) Anzalone, Amin Manna, Amit Saha, Amy Olex, Andrea Zhou, Andrew E. Williams, Andrew Southerland, Andrew T. Girvin, Anita Walden, Anjali A. Sharathkumar, Benjamin Amor, Benjamin Bates, Brian Hendricks, Brijesh Patel, Caleb Alexander, Carolyn Bramante, Cavin Ward-Caviness, Charisse Madlock-Brown, Christine Suver, Christopher Chute, Christopher Dillon, Chunlei Wu, Clare Schmitt, Cliff Takemoto, Dan Housman, Davera Gabriel, David A. Eichmann, Diego Mazzotti, Don Brown, Eilis Boudreau, Elaine Hill, Elizabeth Zampino, Emily Carlson Marti, Emily R. Pfaff, Evan French, Farrukh M Koraishy, Federico Mariona, Fred Prior, George Sokos, Greg Martin, Harold Lehmann, Heidi Spratt, Hemalkumar Mehta, Hongfang Liu, Hythem Sidky, J.W. Awori Hayanga, Jami Pincavitch, Jaylyn Clark, Jeremy Richard Harper, Jessica Islam, Jin Ge, Joel Gagnier, Joel H. Saltz, Joel Saltz, Johanna Loomba, John Buse, Jomol Mathew, Joni L. Rutter, Julie A. McMurry, Justin Guinney, Justin Starren, Karen Crowley, Katie Rebecca Bradwell, Kellie M. Walters, Ken Wilkins, Kenneth R. Gersing, Kenrick Dwain Cato, Kimberly Murray, Kristin Kostka, Lavance Northington, Lee Allan Pyles, Leonie Misquitta, Lesley Cottrell, Lili Portilla, Mariam Deacy, Mark M. Bissell, Marshall Clark, Mary Emmett, Mary Morrison Saltz, Matvey B. Palchuk, Melissa A. Haendel, Meredith Adams, Meredith Temple-O’Connor, Michael G. Kurilla, Michele Morris, Nabeel Qureshi, Nasia Safdar, Nicole Garbarini, Noha Sharafeldin, Ofer Sadan, Patricia A. Francis, Penny Wung Burgoon, Peter Robinson, Philip R.O. Payne, Rafael Fuentes, Randeep Jawa, Rebecca Erwin-Cohen, Rena Patel, Richard A. Moffitt, Richard L. Zhu, Rishi Kamaleswaran, Robert Hurley, Robert T. Miller, Saiju Pyarajan, Sam G. Michael, Samuel Bozzette, Sandeep Mallipattu, Satyanarayana Vedula, Scott Chapman, Shawn T. O’Neil, Soko Setoguchi, Stephanie S. Hong, Steve Johnson, Tellen D. Bennett, Tiffany Callahan, Umit Topaloglu, Usman Sheikh, Valery Gordon, Vignesh Subbian, Warren A. Kibbe, Wenndy Hernandez, Will Beasley, Will Cooper, William Hillegass, Xiaohan Tanner Zhang. Details of contributions available at covid.cd2h.org/core-contributors

The following institutions whose data is released or pending:

Available: Advocate Health Care Network — UL1TR002389: The Institute for Translational Medicine (ITM) •Boston University Medical Campus — UL1TR001430: Boston University Clinical and Translational Science Institute •Brown University — U54GM115677: Advance Clinical Translational Research (Advance-CTR) •Carilion Clinic — UL1TR003015: iTHRIV Integrated Translational health Research Institute of Virginia •Charleston Area Medical Center U54GM104942: West Virginia Clinical and Translational Science Institute (WVCTSI) •Children’s Hospital Colorado — UL1TR002535: Colorado Clinical and Translational Sciences Institute •Columbia University Irving Medical Center — UL1TR001873: Irving Institute for Clinical and Translational Research •Duke University — UL1TR002553: Duke Clinical and Translational Science Institute •George Washington Children’s Research Institute — UL1TR001876: Clinical and Translational Science Institute at Children’s National (CTSA-CN) •George Washington University — UL1TR001876: Clinical and Translational Science Institute at Children’s National (CTSA-CN) •Indiana University School of Medicine — UL1TR002529: Indiana Clinical and Translational Science Institute •Johns Hopkins University — UL1TR003098: Johns Hopkins Institute for Clinical and Translational Research •Loyola Medicine — Loyola University Medical Center •Loyola University Medical Center — UL1TR002389: The Institute for Translational Medicine (ITM) •Maine Medical Center — U54GM115516: Northern New England Clinical & Translational Research (NNE-CTR) Network •Massachusetts General Brigham — UL1TR002541: Harvard Catalyst •Mayo Clinic Rochester UL1TR002377: Mayo Clinic Center for Clinical and Translational Science (CCaTS) •Medical University of South Carolina — UL1TR001450: South Carolina Clinical & Translational Research Institute (SCTR) •Montefiore Medical Center — UL1TR002556: Institute for Clinical and Translational Research at Einstein and Montefiore •Nemours — U54GM104941: Delaware CTR ACCEL Program •NorthShore University HealthSystem — UL1TR002389: The Institute for Translational Medicine (ITM) •Northwestern University at Chicago — UL1TR001422: Northwestern University Clinical and Translational Science Institute (NUCATS) •OCHIN — INV-018455: Bill and Melinda Gates Foundation grant to Sage Bionetworks •Oregon Health & Science University — UL1TR002369: Oregon Clinical and Translational Research Institute •Penn State Health Milton S. Hershey Medical Center — UL1TR002014: Penn State Clinical and Translational Science Institute •Rush University Medical Center — UL1TR002389: The Institute for Translational Medicine (ITM) •Rutgers, The State University of New Jersey — UL1TR003017: New Jersey Alliance for Clinical and Translational Science •Stony Brook University — U24TR002306 •The Ohio State University — UL1TR002733: Center for Clinical and Translational Science •The State University of New York at Buffalo — UL1TR001412: Clinical and Translational Science Institute •The University of Chicago — UL1TR002389: The Institute for Translational Medicine (ITM) •The University of Iowa — UL1TR002537: Institute for Clinical and Translational Science •The University of Miami Leonard M. Miller School of Medicine — UL1TR002736: University of Miami Clinical and Translational Science Institute •The University of Michigan at Ann Arbor — UL1TR002240: Michigan Institute for Clinical and Health Research •The University of Texas Health Science Center at Houston — UL1TR003167: Center for Clinical and Translational Sciences (CCTS) •The University of Texas Medical Branch at Galveston — UL1TR001439: The Institute for Translational Sciences •The University of Utah — UL1TR002538: Uhealth Center for Clinical and Translational Science •Tufts Medical Center — UL1TR002544: Tufts Clinical and Translational Science Institute •Tulane University — UL1TR003096: Center for Clinical and Translational Science •University Medical Center New Orleans — U54GM104940: Louisiana Clinical and Translational Science (LA CaTS) Center •University of Alabama at Birmingham — UL1TR003096: Center for Clinical and Translational Science •University of Arkansas for Medical Sciences — UL1TR003107: UAMS Translational Research Institute •University of Cincinnati — UL1TR001425: Center for Clinical and Translational Science and Training •University of Colorado Denver, Anschutz Medical Campus — UL1TR002535: Colorado Clinical and Translational Sciences Institute •University of Illinois at Chicago — UL1TR002003: UIC Center for Clinical and Translational Science •University of Kansas Medical Center — UL1TR002366: Frontiers: University of Kansas Clinical and Translational Science Institute •University of Kentucky — UL1TR001998: UK Center for Clinical and Translational Science •University of Massachusetts Medical School Worcester — UL1TR001453: The UMass Center for Clinical and Translational Science (UMCCTS) •University of Minnesota — UL1TR002494: Clinical and Translational Science Institute •University of Mississippi Medical Center — U54GM115428: Mississippi Center for Clinical and Translational Research (CCTR) •University of Nebraska Medical Center — U54GM115458: Great Plains IDeA-Clinical & Translational Research •University of North Carolina at Chapel Hill — UL1TR002489: North Carolina Translational and Clinical Science Institute •University of Oklahoma Health Sciences Center — U54GM104938: Oklahoma Clinical and Translational Science Institute (OCTSI) •University of Rochester — UL1TR002001: UR Clinical & Translational Science Institute •University of Southern California — UL1TR001855: The Southern California Clinical and Translational Science Institute (SC CTSI) •University of Vermont — U54GM115516: Northern New England Clinical & Translational Research (NNE-CTR) Network •University of Virginia — UL1TR003015: iTHRIV Integrated Translational health Research Institute of Virginia •University of Washington — UL1TR002319: Institute of Translational Health Sciences •University of Wisconsin-Madison — UL1TR002373: UW Institute for Clinical and Translational Research •Vanderbilt University Medical Center — UL1TR002243: Vanderbilt Institute for Clinical and Translational Research •Virginia Commonwealth University — UL1TR002649: C. Kenneth and Dianne Wright Center for Clinical and Translational Research •Wake Forest University Health Sciences — UL1TR001420: Wake Forest Clinical and Translational Science Institute •Washington University in St. Louis — UL1TR002345: Institute of Clinical and Translational Sciences •Weill Medical College of Cornell University — UL1TR002384: Weill Cornell Medicine Clinical and Translational Science Center •West Virginia University — U54GM104942: West Virginia Clinical and Translational Science Institute (WVCTSI) Submitted: Icahn School of Medicine at Mount Sinai — UL1TR001433: ConduITS Institute for Translational Sciences •The University of Texas Health Science Center at Tyler — UL1TR003167: Center for Clinical and Translational Sciences (CCTS) •University of California, Davis — UL1TR001860: UCDavis Health Clinical and Translational Science Center •University of California, Irvine — UL1TR001414: The UC Irvine Institute for Clinical and Translational Science (ICTS) •University of California, Los Angeles — UL1TR001881: UCLA Clinical Translational Science Institute •University of California, San Diego — UL1TR001442: Altman Clinical and Translational Research Institute •University of California, San Francisco — UL1TR001872: UCSF Clinical and Translational Science Institute Pending: Arkansas Children’s Hospital — UL1TR003107: UAMS Translational Research Institute •Baylor College of Medicine — None (Voluntary) •Children’s Hospital of Philadelphia UL1TR001878: Institute for Translational Medicine and Therapeutics •Cincinnati Children’s Hospital Medical Center — UL1TR001425: Center for Clinical and Translational Science and Training •Emory University — UL1TR002378: Georgia Clinical and Translational Science Alliance •HonorHealth — None (Voluntary) •Loyola University Chicago — UL1TR002389: The Institute for Translational Medicine (ITM) •Medical College of Wisconsin — UL1TR001436: Clinical and Translational Science Institute of Southeast Wisconsin •MedStar Health Research Institute — UL1TR001409: The Georgetown-Howard Universities Center for Clinical and Translational Science (GHUCCTS) •MetroHealth — None (Voluntary) •Montana State University — U54GM115371: American Indian/Alaska Native CTR •NYU Langone Medical Center — UL1TR001445: Langone Health’s Clinical and Translational Science Institute •Ochsner Medical Center — U54GM104940: Louisiana Clinical and Translational Science (LA CaTS) Center •Regenstrief Institute — UL1TR002529: Indiana Clinical and Translational Science Institute •Sanford Research — None (Voluntary) •Stanford University — UL1TR003142: Spectrum: The Stanford Center for Clinical and Translational Research and Education •The Rockefeller University — UL1TR001866: Center for Clinical and Translational Science •The Scripps Research Institute — UL1TR002550: Scripps Research Translational Institute •University of Florida — UL1TR001427: UF Clinical and Translational Science Institute University of New Mexico Health Sciences Center — UL1TR001449: University of New Mexico Clinical and Translational Science Center •University of Texas Health Science Center at San Antonio — UL1TR002645: Institute for Integration of Medicine and Science •Yale New Haven Hospital — UL1TR001863: Yale Center for Clinical Investigation

Authorship was determined using ICMJE recommendations.

## Conflict of Interest

The authors declare no conflicts of interest.

## Supplemental Figures S1-S3

**Figure S1.**
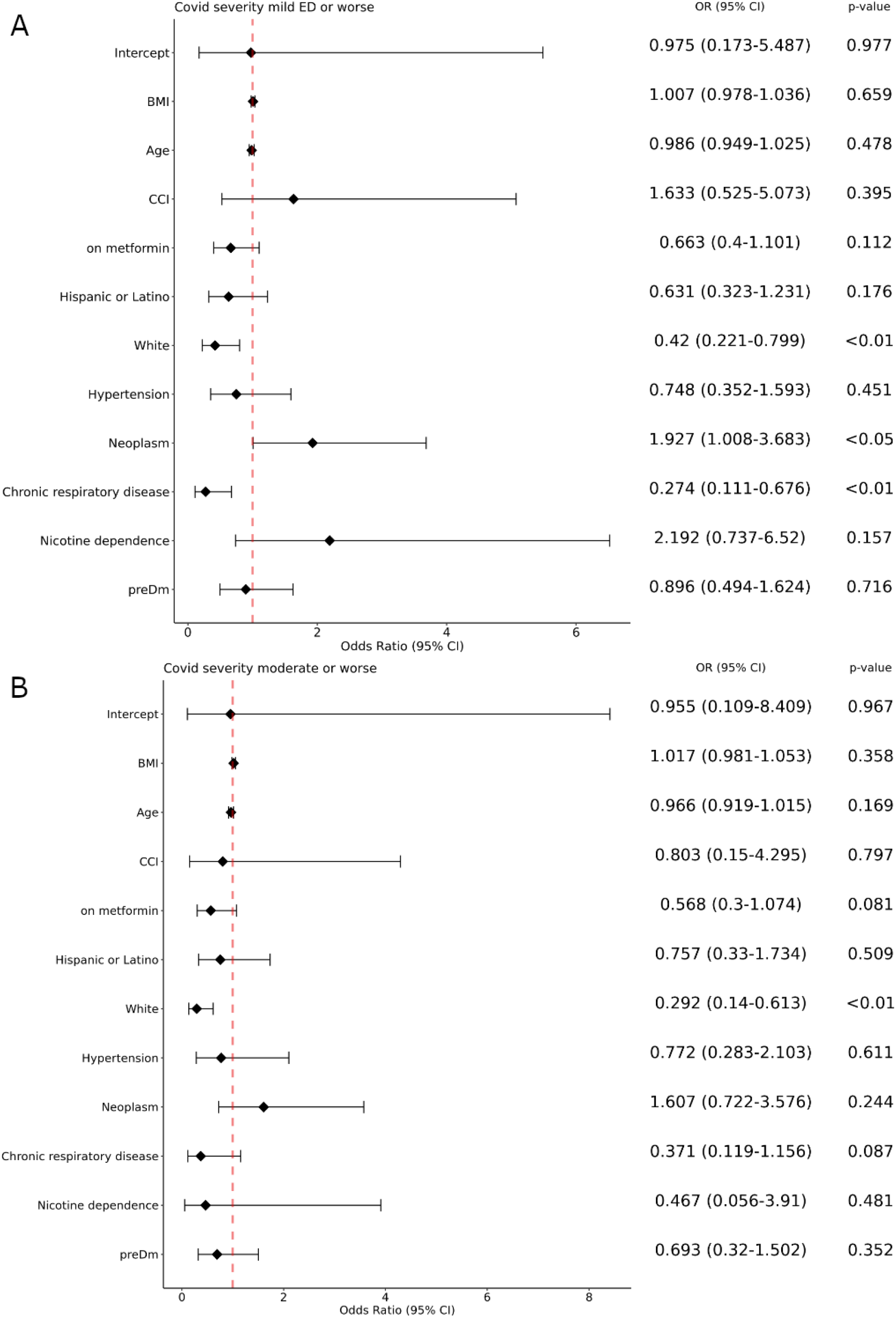
Forest plots for the PCOS cohort. A: Mild vs Mild ED and worse; B: Mild and Mild ED vs Moderate and worse.

**Supplementary Figure S2.**
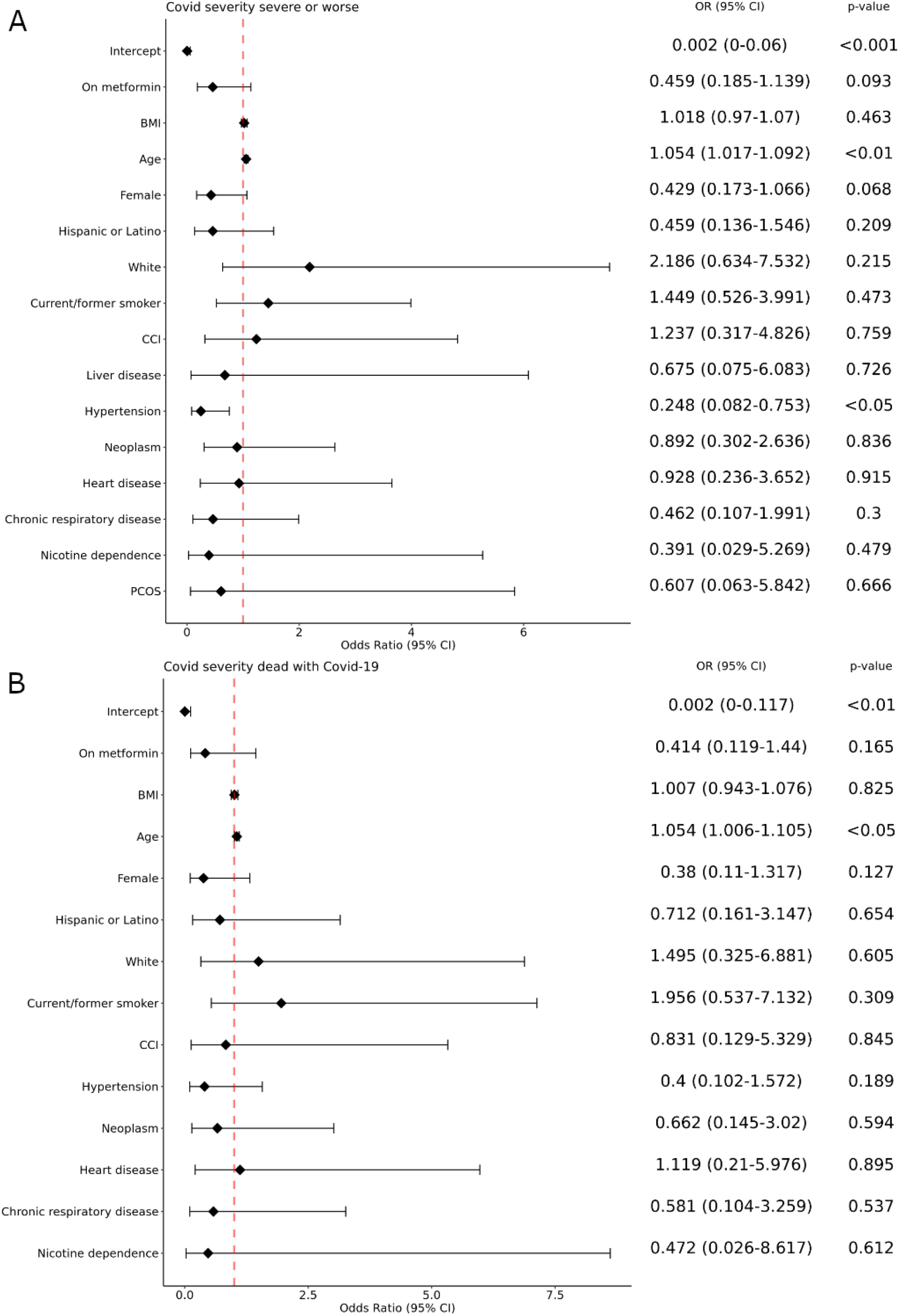
Forest plots for the prediabetes cohort. A: Mild, mild ED, and moderate vs severe and worse, B: Mild, mild ED, moderate, and severe vs death with COVID-19

**Supplementary Figure S3.**
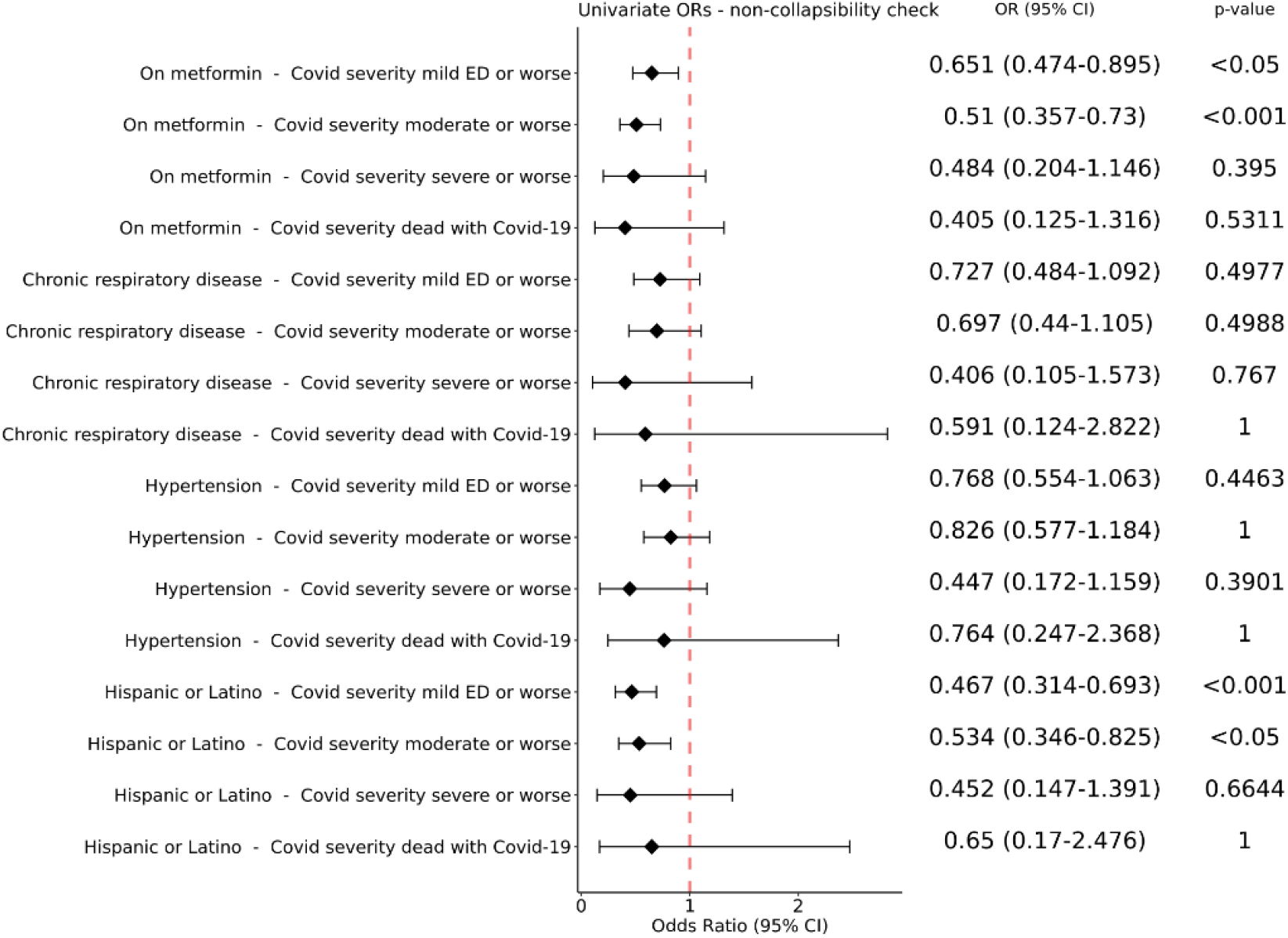
Forest Plots for univariate odds ratios computed to assess the non-collapsibility effect for chronic respiratory disease. According to Schuster et al.,^50^ non-collapsibility effects on a covariate can be estimated by comparing the OR obtained by univariate LR on the variable and the OR obtained in the multivariate analysis. To check whether the OR for chronic-respiratory disease is affected by the non-collapsibility effect we therefore run an univariate LR for each outcome. The forest plot reports the results obtained when univariate LRs were applied to either drug usage (“on metformin”) or chronic respiratory disease; the ORs for drug usage are comparable to those obtained by multivariate LR (Fig. 4a, 4b) showing the reliability of the estimate. On the other hand, the remarkable difference in the ORs estimated for chronic respiratory disease confirm the non-collapsibility effect for this variable.

## Notes

### Competing Interest Statement

The authors have declared no competing interest.

### Author Declarations

The analyses described in this publication were conducted with data or tools accessed through the NCATS N3C Data Enclave covid.cd2h.org/enclave and supported by CD2H - The National COVID Cohort Collaborative (N3C) IDeA CTR Collaboration 3U24TR002306-04S2 NCATS U24 TR002306, DUR: RP-7BE1AC. This research was possible because of the patients whose information is included within the data from participating organizations (covid.cd2h.org/dtas) and the organizations and scientists (covid.cd2h.org/duas) who have contributed to the on-going development of this community resource. The N3C data transfer to NCATS is performed under a Johns Hopkins University Reliance Protocol #IRB00249128 or individual site agreements with NIH. The N3C Data Enclave is managed under the authority of the NIH; information can be found at https://ncats.nih.gov/n3c/resources.

